# Predictors of successful weaning from Veno-Arterial Extracorporeal Membrane Oxygenation (V-A ECMO): A Systematic Review and Meta-analysis

**DOI:** 10.1101/2024.08.30.24312815

**Authors:** Henry R. Hsu, Praba Sekhar, Jahnavi Grover, David H. Tian, Ciaran Downey, Ben Maudlin, Chathuri Dissanayake, Mark Dennis

## Abstract

**Background:** Venoarterial extracorporeal membrane oxygenation (V-A ECMO) use to support patients in cardiac failure is increasing. Despite this increased use, predicting successful weaning from ECMO can be challenging, no uniform guidelines on weaning exist. Therefore, we completed a systematic review to evaluate prognostic factors that predict successful weaning from V-A ECMO.

**Methods:** Following the PRIMSA guidelines, a systematic literature search of Medline, Embase, SCOPUS and CENTRAL identified original research studies of patients requiring V-A ECMO where weaning was attempted. Data was collected on demographic factors and weaning protocol, biomarkers, haemodynamic, echocardiographic factors for the successfully weaned (SW) and not successfully weaned (NSW) groups. Two investigators reviewed studies for relevance, extracted data, and assessed risk of bias using the ROBINS-I tool. The study was registered on the international prospective register of systematic reviews (PROSPERO ID# CRD42022366153).

**Results:** 1219 records were screened, of which 20 studies were deemed sufficient to be included in the statistical analysis based on pre-specified criteria. Factors associated with successful weaning were higher left ventricular ejection fraction (LVEF) (MD 9.0, 95% CI 4.1 – 13.8; p<0.001) and left ventricular outflow tract velocity time integral (LVOT VTI) at time of weaning, (MD 1.35, 95% CI 0.28 – 2.40 lactate at admission (MD -2.5, 95%CI -3.8 – -1.1, p<0.001;), and CK-MB at admission (MD -4.11, 95%CI -6.6 to -1.6, p=0.001). Critical appraisal demonstrated moderate-high risk of bias owing to confounding and low sample sizes.

**Conclusion:** In patients on V-A ECMO support being assessed for weaning multi-parametric assessment is required. Moderate-high heterogeneity and low sample sizes warrant higher-quality studies to help guide decisions to wean patients from V-A ECMO.

## INTRODUCTION

Veno-arterial (V-A) extracorporeal membrane oxygenation (ECMO) is increasingly being used (1, 2) in acute cardiac failure to provide end-organ perfusion whilst definitive treatment, myocardial recover occurs or bridge to left ventricular assist device (LVAD) or heart transplant is completed.

Complications whilst on V-A ECMO support are common and effect mortality and increase with duration of support(3). Therefore, minimising duration of V-A ECMO support, where possible, is sought. However, premature withdrawal of V-A ECMO support, may result in recurrence of cardiogenic shock and effect recovering organs. Minimizing complications associated with device support with the potential for hemodynamic deterioration if support is prematurely discontinued can be challenging.

The definition of successful V-A ECMO weaning has been proposed as when a patient survives for longer than 48 hours after ECMO explantation, with more, recent definitions as those having ECMO removed and not requiring further mechanical support because of recurring cardiogenic shock over the following 30 days(4, 5). Depending on the definition the proportion of V-A ECMO patients successfully weaned ranged between 30-75%(4–9).

A variety of clinical, haemodynamic, biochemical and echocardiographic variables have been proposed and used to guide clinical improvement and readiness to wean(10). However, criteria and variables have not been completely reviewed to ascertain effectiveness(11) and meta-analyses as yet not completed. Therefore, we systematically reviewed a broad range of biomarkers, haemodynamic, echocardiographic and scoring systems to predict successful weaning from V-A ECMO.

## METHODS

The study was conducted as previously outlined in our registered and published protocol (PROSPERO ID# CRD42022366153) and in accordance with the PRISMA (Preferred Reporting Items for Systematic Reviews and Meta-Analyses) guidelines(12). Ethics approval and patient consent were not required.

### Search strategy

The search strategy is detailed in Supplementary Material. A comprehensive search of three electronic databases (Medline, Embase, SCOPUS and CENTRAL) was conducted in October 2022, which were re-run in December 2023 and March 2024 prior to final analysis and further studies retrieved for inclusion. Appropriate Boolean operators were used to combine search terms that included V-A ECMO, ECMO, extra-corporeal life support, weaning, decannulation and ECLS. The reference lists of all included studies were also reviewed to identify any additional articles, and duplicate articles were removed. Studies that were not primarily in the English language were included if they were accompanied by an English translation. There were no limitations on the publication period.

### Study characteristics

Inclusion criteria allowed for randomized controlled trials, cohort studies, case series and conference abstracts that (1) considered adult or paediatric populations (2) involved patients who were on V-A ECMO and (3) there was an attempt to de-cannulate/wean from ECMO. Studies using ECMO as a bridge to ventricular assist device or heart transplant were excluded. Case series were included if >5 patients. Studies had to report associations between variables within the study and weaning success. Review publications, grey literature, non-English language publications, editorials, comments, letters to the editor and animal studies were excluded. Studies only assessing baseline variables with weaning success were excluded.

### Study Selection

Title and abstract screening were conducted by independent investigators (P.S. or H.H. or C.D.). Likewise, full-text screening was performed by two independent investigators (P.S. or H.H. or C.D.). All conflicts were resolved by a third, senior investigator (M.D.). The systematic review platform Covidence (www.covidence.org; Veritas Health Innovation, Melbourne, Australia) was used to facilitate the screening process. Publications found to fulfil eligibility criteria underwent data extraction.

### Data extraction

Data was extracted from studies by two independent reviewers (P.S. or H.H. or C.D. or J.G.) using Microsoft Excel. Extracted variables included but not limited to patient demographics, weaning protocol, successful weaning definition, weaning success, various prognostic factors including biomarkers, haemodynamic, echocardiographic and scoring systems. The primary outcome was weaning success defined survival post removal of mechanical circulatory support and not requiring ventricular assist device or heart transplant. Meta-analysis was planned of predictors as appropriate. Missing data was reported as not reported. Authors were attempted to be contacted for further or missing data via email.

### Evaluation of risk of bias

Critical appraisal of the risk of bias for individual studies was conducted using the ROBINS-I Tool (Risk of Bias in Non-Randomized Studies - of Interventions)(13). Each included study was scored by two independent investigators (P.S. or J.G. or H.H.). Any discrepancies between the two reviewers were resolved by discussion and mutual agreement. Studies of poor-quality following risk of bias assessment were not be excluded from being included in our synthesis. Where a poor-quality study has contributed to a synthesized effect estimate, we explored the impact of study quality by performing sensitivity analysis by removing the poor-quality study to observe the impact that bias has had on the synthesized effect.

### Statistical analysis

Meta-analysis was completed as per the Cochrane Handbook for Systematic Reviews of Interventions when the outcomes were reported by two or more trials (14). Statistical analysis was performed using Review Manager (version 5.3, The Cochrane Collaboration, Oxford, UK). For continuous outcomes, mean, standard deviation (SD) and sample size were extracted from each of the groups. Where studies reported median and ranges or interquartile range, derived mean and standard deviation as described by Wan et al. were calculated. (15). Mean differences with 95% confidence interval (CI) were used for continuous outcomes. An inverse variance method was applied for mean difference. Heterogeneity was assessed using I2 statistics and values between 50% and 90% were considered to represent substantial heterogeneity. A random effects meta-analysis model and exploring heterogeneity with sensitivity and subgroup analysis were applied where appropriate. Categorisation of reported risk factors of successful weaning from studies that reported multivariable adjustment was completed.

## RESULTS

### Systematic search and study selection

The search strategy of relevant references yielded a total of 2199 references (Figure 1). After the removal of 980 duplicates, the remaining 1219 references were screened by title and abstract. A total of 62 publications were deemed to be eligible for full-text screening, of which 28 studies were excluded with reasons. A total of 34 articles were included in the final analysis, of which 20 studies were deemed sufficient to be included in the statistical analysis. Risk of bias assessment is summarised in Figure 2. The remaining 14 studies examined differing prognostic predictors that were not able to be meta-analysed together. The sample sizes ranged from 12 to 265 patients, with a pooled sample size of 1903 patients. Twenty-six publications were retrospective cohort studies, seven were prospective cohort studies, and one not recorded. Cardiogenic shock was the primary indication for V-A ECMO in ten publications, myocarditis in two, cardiomyopathy in two, cardiac arrest in four, pulmonary embolism in one, post cardiac surgery in two, congenital heart disease in one, acute respiratory distress syndrome in one, and eleven not recorded. Geographically, fourteen publications were from Asia, eight were from North America, eleven were from Europe, and one was from the Middle East. A summary of the baseline characteristics of included papers and clinical variables is provided in Table 1.

**Figure 1:**
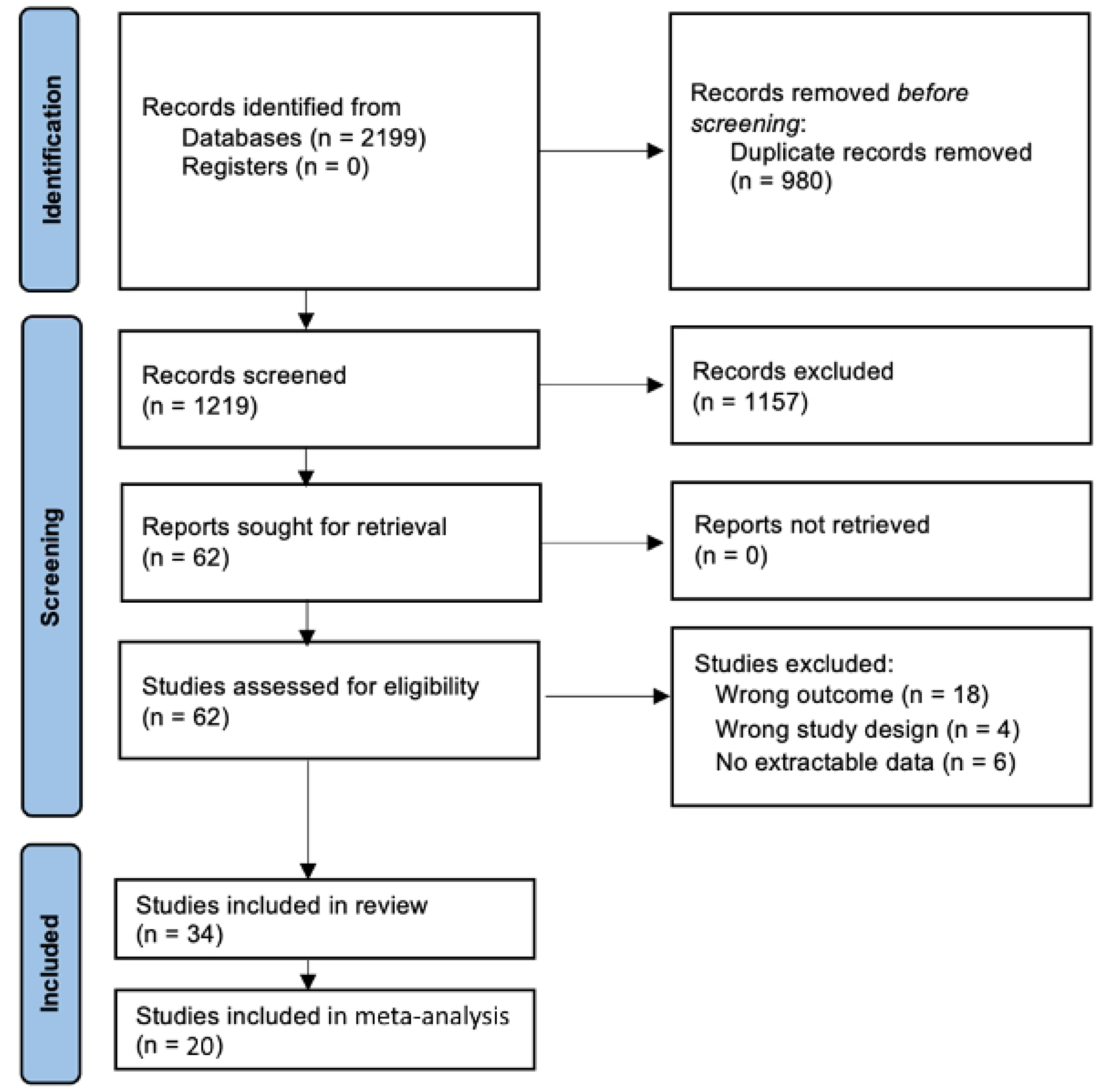
PRISMA flow diagram illustrating the number of studies identified by the search and the stages in which they were chosen and eliminated.

**Figure 2:**
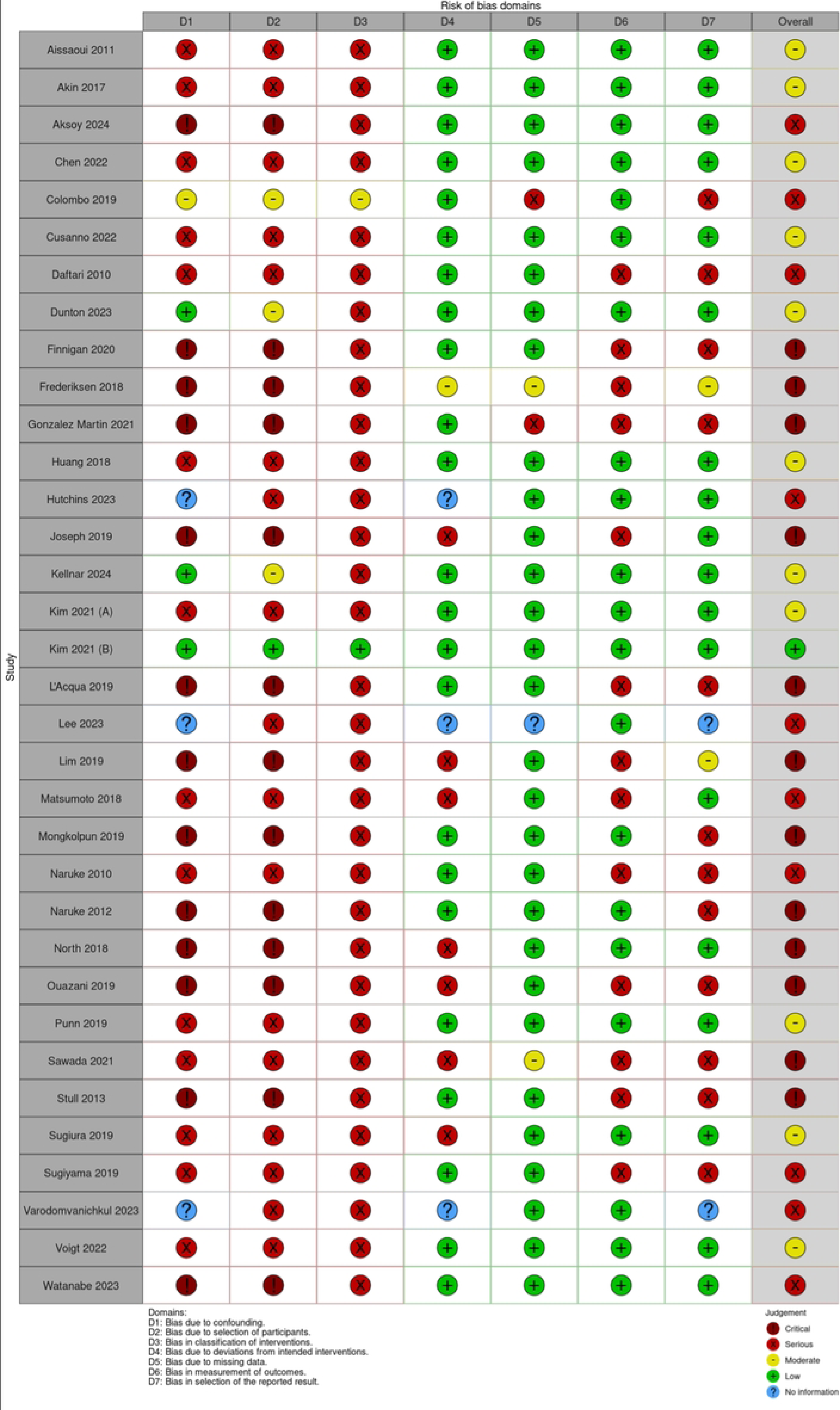
Risk of bias of the included studies (ROBINS-I)

**Table 1:**
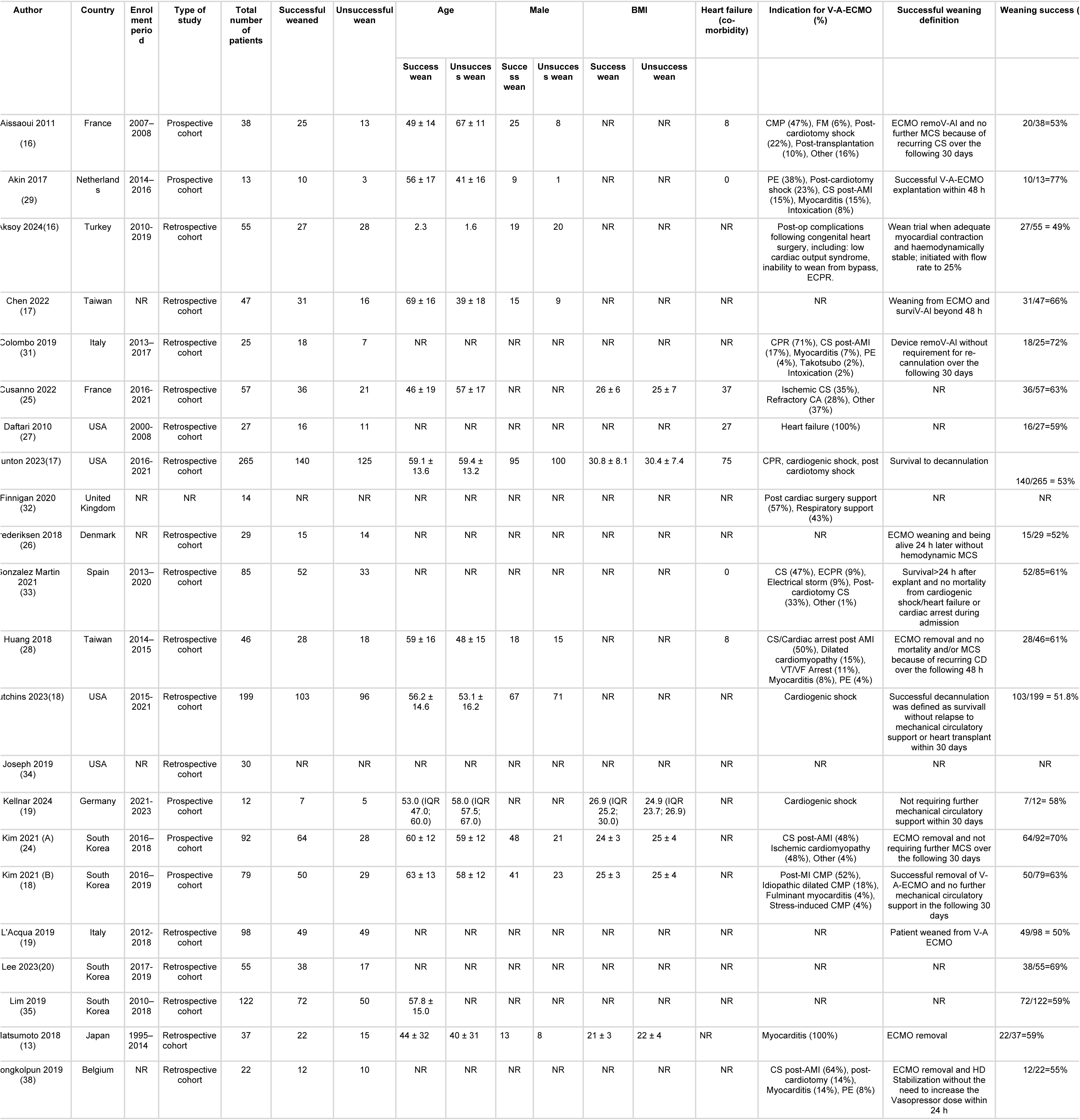
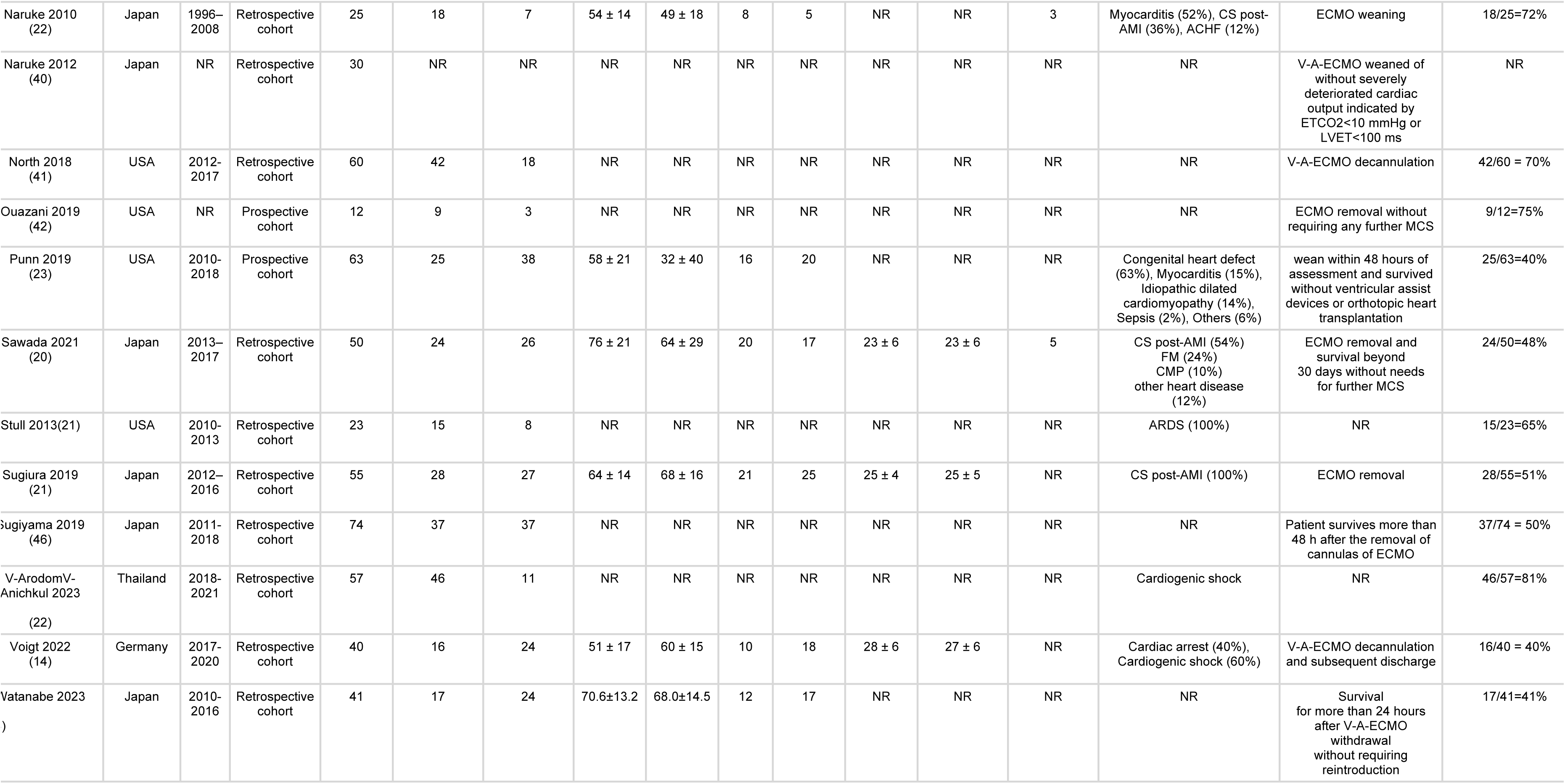
Characteristics of included studies.

ACHF acute on chronic heart failure, AMI acute myocardial infarction, ARDS acute respiratory distress syndrome, CA Cardiac arrest, CMP cardiomyopathy, CPR cardiopulmonary resuscitation, CS cardiogenic shock, ECMO extracorporeal membranous oxygenation, FM fulminant myocarditis, MCS mechanical cardiac support, NR not reported, PE pulmonary embolism, VF ventricular fibrillation, VT ventricular tachycardia

### Biomarkers

Markers of organ damage were inversely associated with weaning success – Figure 3. Lower levels of creatinine kinase (CK-MB) (MD -4.1, 95%CI -6.6 – -1.6, p=0.001; I2 = 24%), lactate at admission (MD -2.5, 95%CI -3.8 – -1.1, p<0.001; I2 =85%), and lower levels of alanine aminotransferase (ALT) (MD -36.7, 95%CI -65.5 – 7.9, P=0.01; I2 = 0%) at the time of weaning were associated with weaning success. Too few studies reported NT-ProBNP or Troponin to enable analysis.

**Figure 3.**
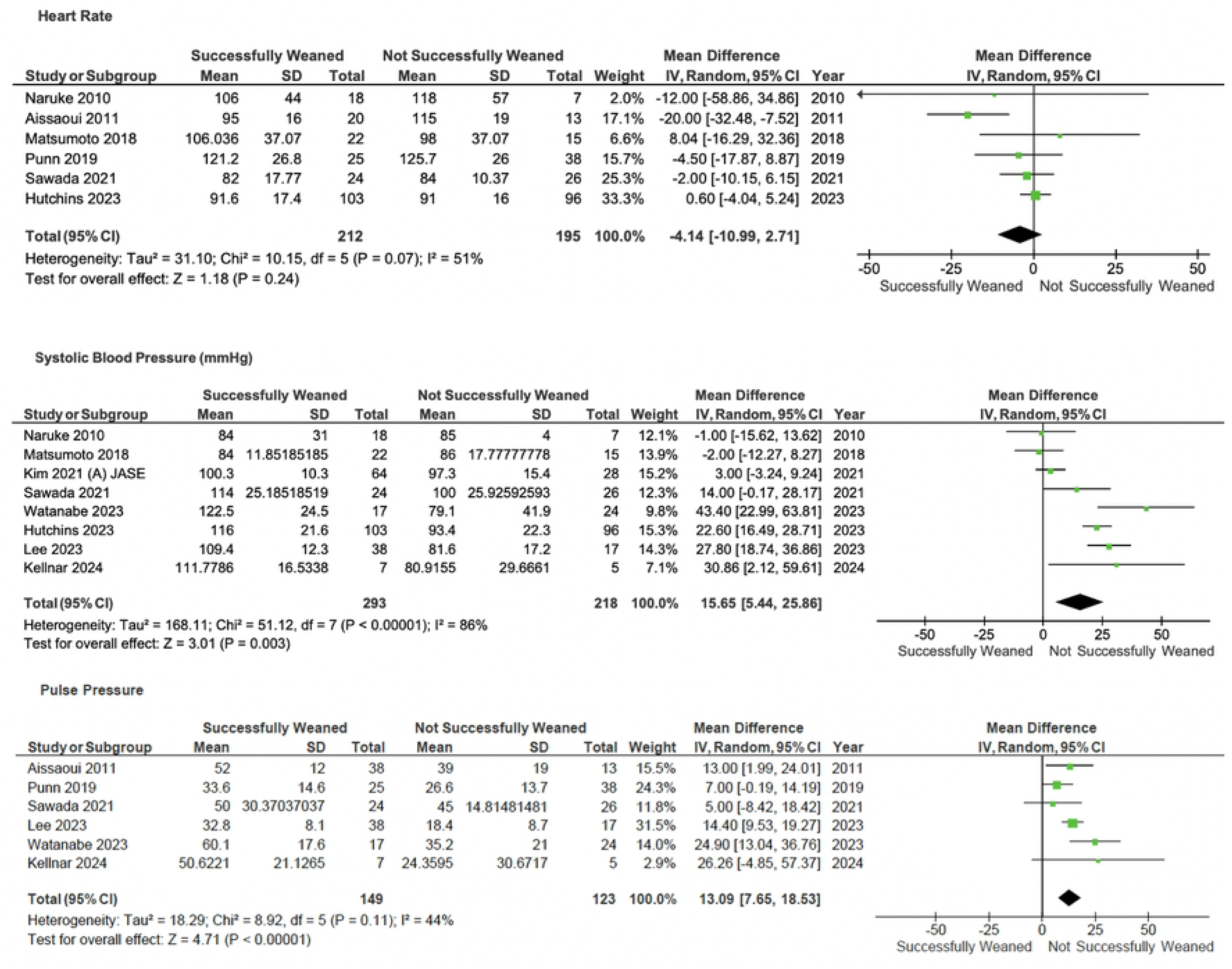
Forrest plot of comparison of haemodynamic parameters on V-A ECMO Weaning.

### Haemodynamics

Patients with higher pulse pressure (MD 13.1, 95%CI 7.7 – 18.5, p<0.001; I2 = 44%) and systolic blood pressure (MD 15.7, 95%CI 5.4 – 25.9, p<0.001; I2 = 86%) were associated with weaning success – Figure 4.

**Figure 4.**
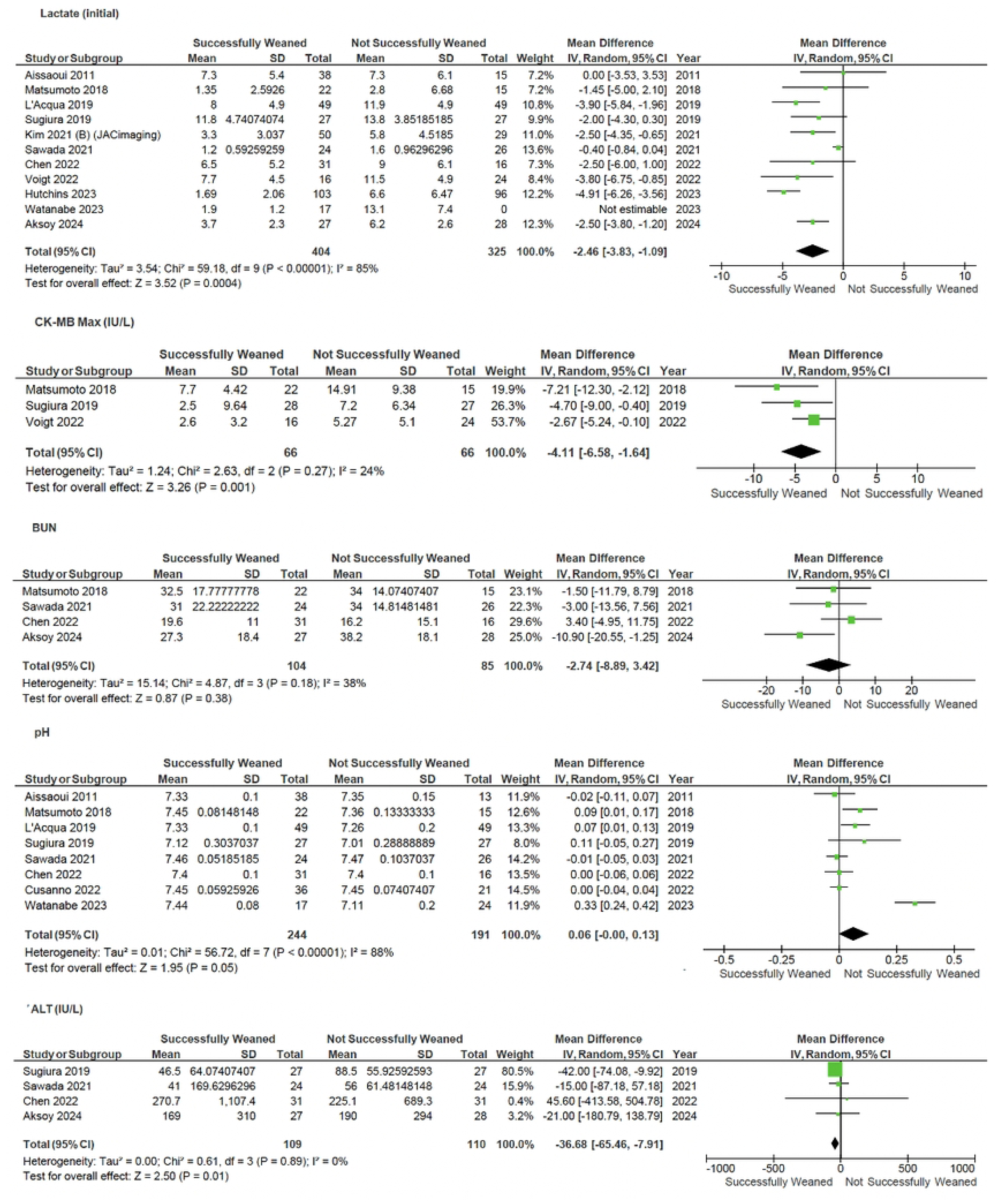
Forrest plot of comparison of laboratory parameters on V-A ECMO.

### Echocardiography

Patients with higher left ventricular ejection fraction (LVEF) at time of weaning (MD 9.0, 95% CI 4.1 – 13.8; p<0.001; I2 = 85%), left ventricular outflow tract velocity time integral (LVOT VTI) (MD 1.35, 95% CI 0.28 – 2.40, p=0.01; I2 = 0%), E/Ea (MD -2.72, 95% CI -4.45 - -0.98, p=0.002; I2 = 29%) were associated with weaning success – Figure 5.

**Figure 5.**
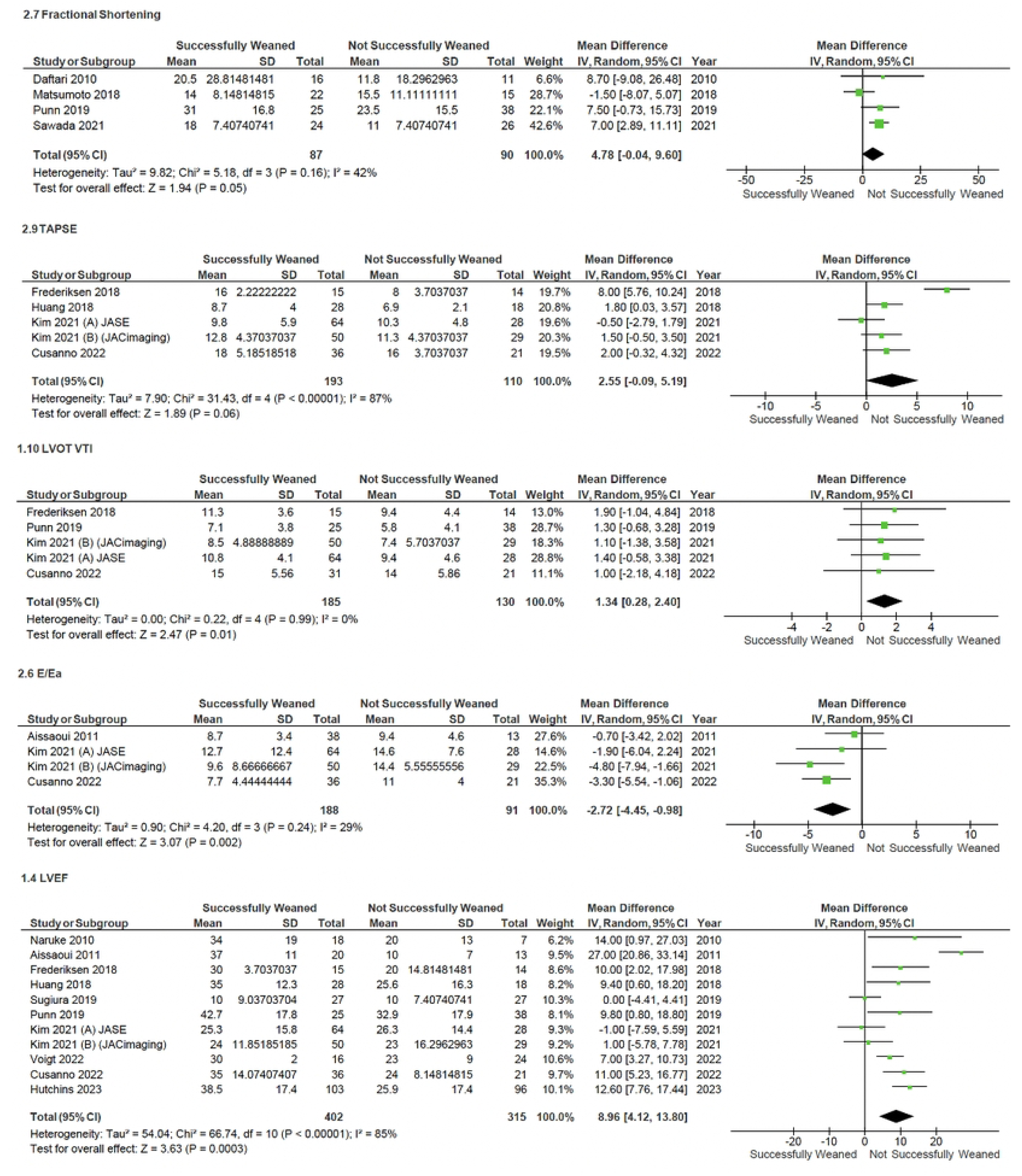
Forest plot of the comparison of different echocardiographic parameters on V-A ECMO weaning.

### Multi-variable adjusted: Risk predictors

Thirteen studies provided multi-variable adjusted analysis to identify predictors of successful weaning. Covariates tested varied widely between studies; only lack of renal failure or CRRT during ECMO, and post-weaning lactate, reported by more than one study as predictors of successful ECMO weaning – Table 2.

**Table 2.**
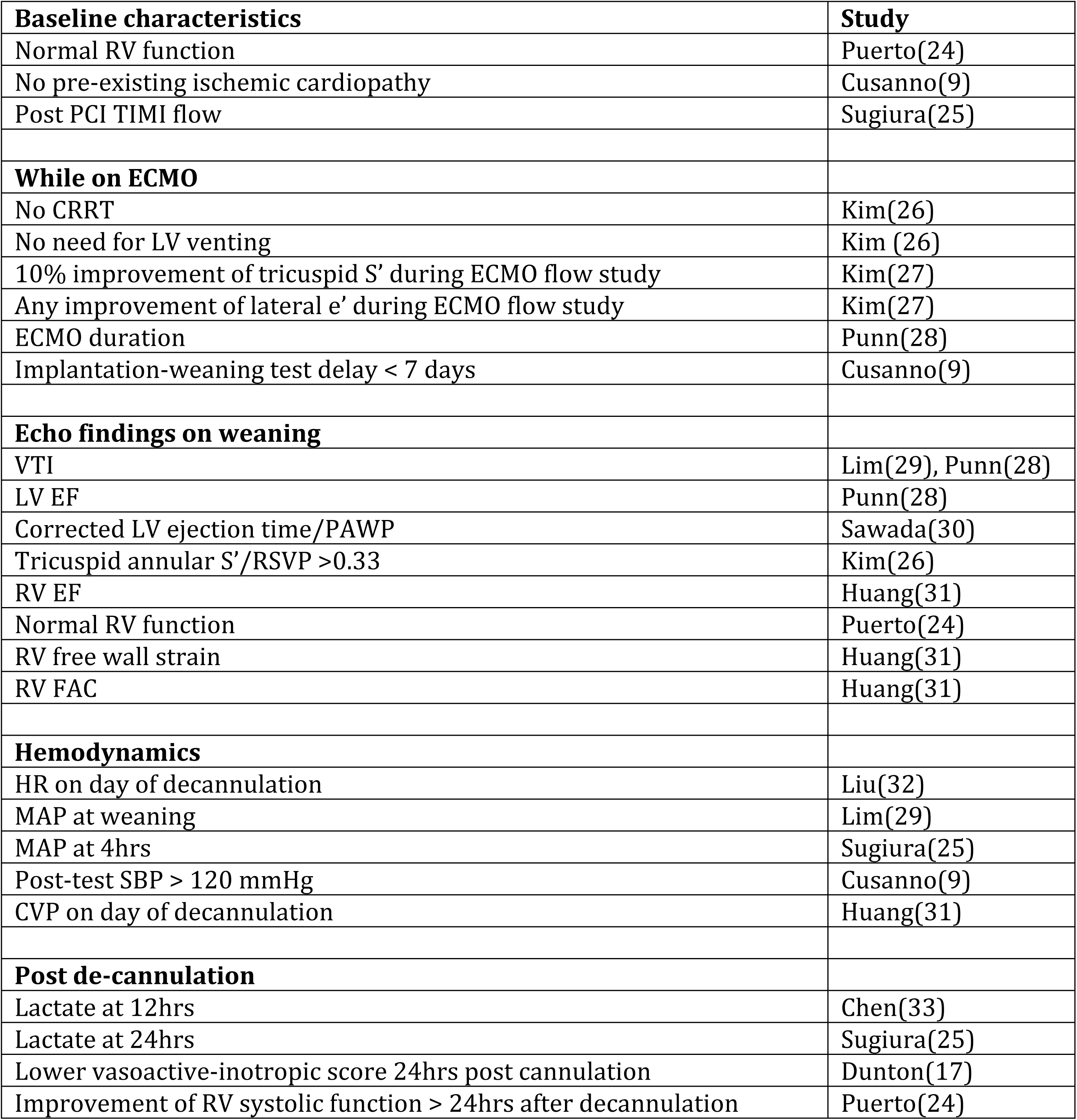
Risk factors predictive of successful ECMO weaning from multi-variable logistic regression models. CRRT, continuous renal replacement therapy; CVP, central venous pressure; ECMO, extracorporeal membrane oxygenation; EF, ejection fraction; FAC, fractional area change; HR, heart rate; LV, left ventricle; **MAP**, mean arterial pressure; PAWP, pulmonary artery wedge pressure; PCI, percutaneous coronary intervention; RV, right ventricle; SBP, systolic blood pressure; TIMI, Thrombolysis in Myocardial Infarction score; VTI, velocity time integral.

### Study quality

A total of 34 publications were eligible for quality assessment, of which 12 publications were of poor quality, i.e. ‘critical’ risk of bias (Figure 2). Many studies did not provide detailed protocols, limiting methodological assessment, appraisal of the confounding effect of intervention and bias in selection of participants into the study.

## DISCUSSION

In this systematic review of predictors of V-A ECMO weaning success 34 predominantly small observational, studies were identified. On pooled analysis, lower levels of biochemical markers of end-organ perfusion or injury (lactate, CK-MB and ALT), haemodynamic (pulse pressure and systolic blood pressure) and echocardiographic indicators of myocardial function (LVEF, LVOT VTI, E/Ea) were associated with successful weaning form V-A ECMO.

To our knowledge this review is only the second to systematically assess predictors of V-A ECMO wean success. The first, in adult patients with specifically cardiogenic shock or cardiac arrest identified similar results to our review, with lower creatine kinase and lactate levels, and higher LVEF being predictors for successful weaning from V-A-ECMO (10) Other, non-systematic, reviews have reported also reported lower creatine kinase and lactate levels, and higher LVEF and LVOT VTI being predictors for successful weaning from V-A-ECMO (34). Several other V-variables used in clinical practice (9) i.e. Troponin, NT-ProBNP, RV to PA coupling indices were not identified owing to limited numbers of studies and patients reported with these.

Despite significant heterogeneity, small sample sizes and a significant risk of bias, there are some conclusions that can be drawn from this review and the available literature. First, determination of likely weaning success, should consider multiple variables and not be focussed on one individual predictor. Factors associated with success (or failure), were present across clinical, biochemical, haemodynamic and echocardiographic parameters and clinicians should avoid relying on one variable over the complete picture of the patient. Second, initial severity of illness (e.g. lactate), markers of end-organ perfusion, and then recovery of such are important considerations in attempting to wean (25, 35, 36). Further, absolute cut offs for specific variables e.g. LVEF or LVOT VTI to predict weaning success vary between studies, are based empirical clinical weaning protocols (34, 37) and therefore cannot yet be elucidated. Overall restitution and improvement of the overall clinical state of the patient as well as cardiac function is likely key to successful weaning rather than a specific variable or level of a variable.

Formal weaning or “ramp” studies that assess haemodynamic and echocardiographic changes to alterations to ECMO flows protocols are recommended (38) but as yet no standardised protocols exist, are only variably reported in ECMO trials, and are not formally assessed in this systematic review. However, they are critical tools to assess the response of cardiac function to reduction, and then removal, of mechanical circulatory support (9, 39). Future prospective clinical trials should publish weaning strategies and protocols to enable further assessment and comparison of strategies.

### Limitations

Our review is limited by the lack of large high-quality trial, with all included studies consisting of observational studies with small sample sizes and these small trials were used to investigate widely varying interventions amongst this population group, often performed without covariate adjustment. However, we completed a comprehensive review of the literature including all commonly used variables for V-A ECMO weaning. Micro-circulation indices were not assessed but are not in uniform clinical practice which was our focus.

## CONCLUSIONS

In patients requiring V-A ECMO support, multiple biochemical, haemodynamic and echocardiographic parameters of recovery, rather than a single variable should be used to guide appropriateness for weaning. Further larger studies are required to determine optimal weaning strategies.

## Data Availability

All relevant data are within the manuscript and its Supporting Information files.

## AUTHOR CONTRIBUTIONS

Praba Sekhar, Henry Hsu and Mark Dennis conceived and designed the project. Praba Sekhar, Henry Hsu, Ciaran Downey and Jahnavi Grover conducted the systematic review with guidance from Mark Dennis. Praba Sekhar, Jahnavi Grover and Mark Dennis drafted the manuscript. Chathuri Dissanayake and Ben Maudlin provided critical clinical input and advice with the manuscript. All authors read, edited, and approved the final manuscript prior to submission.

## ACKNOWLEDGMENTS

Although this project received no specific funding, one author is currently supported by funding from the National Heart Foundation of Australia and The University of Sydney (M.D.).

## CONFLICT OF INTEREST

None of the authors has any conflict of interest to disclose.

## Notes

### Competing Interest Statement

The authors have declared no competing interest.

### Funding Statement

The author(s) received no specific funding for this work.

