## Supplementary Material for "Predictors of successful weaning from Veno-Arterial Extracorporeal Membrane Oxygenation (V-A ECMO): A Systematic Review and Meta-analysis"

Search Strategy

Dates Run: 10th Oct 2022 (rerun - Dec 2023, Mar 2024)

**Ovid - Embase, Medline, CENTRAL**

| #1 | Extracorporeal Membrane Oxygenation/ | 47297 |
| --- | --- | --- |
| #2 | ECMO.mp. | 36920 |
| #3 | wean.mp. | 142926 |
| #4 | de-cannulat*.mp. | 126 |
| #5 | decannulat*.mp. | 8157 |
| #6 | 1 or 2 | 61919 |
| #7 | 3 or 4 or 5 | 149897 |
| #8 | Prognosis/ | 1242751 |
| #9 | Predictive Value of Tests/ | 378943 |
| #10 | Predict*.mp. | 4811427 |
| #11 | prognos*.mp. | 2439697 |
| #12 | 8 or 9 or 10 or 11 | 6570094 |
| #13 | 6 and 7 and 12 | 1719 |

**Scopus**

| #1 | (INDEXTERMS("extracorporeal membrane oxygenation") OR TITLE-ABS-KEY(ECMO)) AND (TITLE-ABS-KEY(wean*) OR TITLE-ABS-KEY(de-cannulat*) OR TITLE-ABS-KEY(decannulat*)) AND (INDEXTERMS(Prognosis) OR INDEXTERMS("Predictive Value of Tests") OR TITLE-ABS-KEY(Predict*) OR TITLE-ABS-KEY(prognos*)) | 480 |
| --- | --- | --- |
